# Transforming Estonian health data to the Observational Medical Outcomes Partnership (OMOP) Common Data Model: lessons learned

**DOI:** 10.1101/2023.02.16.23285697

**Authors:** Marek Oja, Sirli Tamm, Kerli Mooses, Maarja Pajusalu, Harry-Anton Talvik, Anne Ott, Marianna Laht, Maria Malk, Marcus Lõo, Johannes Holm, Markus Haug, Hendrik Šuvalov, Dage Särg, Jaak Vilo, Sven Laur, Raivo Kolde, Sulev Reisberg

## Abstract

**Objective:** To describe the reusable transformation process of electronic health records (EHR), claims, and prescriptions data into Observational Medical Outcome Partnership (OMOP) common data model (CDM), together with challenges faced and solutions implemented.

**Materials and Methods:** We used Estonian national health databases that store almost all residents’ claims, prescriptions, and EHR records. To develop and demonstrate the transformation process of Estonian health data to OMOP CDM, we used a 10% random sample of the Estonian population (n = 150,824 patients) from 2012-2019. For the sample, complete information from all three databases was converted to OMOP CDM version 5.3. The validation was performed using open-source tools.

**Results:** In total, we transformed over 100 million entries to standard concepts using standard OMOP vocabularies with the average mapping rate 95%. For conditions, observations, drugs, and measurements, the mapping rate was over 90%. In most cases, SNOMED Clinical Terms were used as the target vocabulary.

**Discussion:** During the transformation process, we encountered several challenges, which are described in detail with concrete examples and solutions.

**Conclusion:** For a representative 10% random sample, we successfully transferred complete records from three national health databases to OMOP CDM and created a reusable transformation process. Our work helps future researchers to transform linked databases into OMOP CDM more efficiently, ultimately leading to better real-world evidence.

## BACKGROUND AND SIGNIFICANCE

While randomized controlled trials are the gold standard for causative clinical studies, generating real-world evidence (RWE) from routinely collected real-world health data (RWD) has gained more and more attention in recent years as it provides information about a broader patient population in a less controlled environment when compared to the clinical trials, and it better reflects what is actually happening in the clinical practice[1–3]. RWD can be used for a large variety of studies – for example, for the scientific evaluation of usage, effects, potential benefits, and risks of a medical product[3], drug adherence, health and treatment patterns, treatment guidelines[4].

To generate high-quality RWE, diverse RWD from different healthcare settings and geographical locations integrated into data networks are needed[1]. Still, linking different real-world datasets and conducting multi-database studies to produce high-quality RWE is a challenging task. It has been shown that many of the problems could be solved by using a single common data model for all datasets[5–7]. This would improve not only the quality of the outcome but also their acceptability in decision-making[6]. In recent years, there has been an increasing interest in transferring health data to the Observational Medical Outcome Partnership (OMOP) common data model (CDM)[8,9], which offers a standardized vocabulary and structure, improving the interoperability between databases. Moreover, several open-source software solutions have been developed to support the transformation and analysis process[10]. This all supports the transformation process of health data to OMOP and could be considered one key reason why OMOP has become increasingly popular.

Previous research has described the successful transformation of data to OMOP CDM, which originate from different sources like biobanks[11], national databases, and registries[12–14], hospital databases[15–18], questionnaires[19], cohort studies[20–22]. Some studies focus on specific conditions or some part of a database[12,13,16,17,19,21] while others transfer whole databases with different diagnoses, drug adherence or health care procedures[11,15,20]. Despite the existing research, it has been stressed that continued sharing of experiences, methodologies, and challenges of the data transformation process to OMOP is needed as it helps to develop the transformation process and foster collaboration[21,23].

Today OHDSI network includes more than 453 databases mapped to the OMOP CDM[10], and an initiative to establish a federated network of OMOP healthcare datasets across Europe has been coordinated and partly funded in the European Health Data & Evidence Network (EHDEN) project[24]. However, the geographic distribution of OMOP datasets is uneven as the real-world datasets from Eastern European countries are much less represented than Western countries, leading to gaps in real-world evidence from these regions[25]. In addition, the number of datasets that contain data from several healthcare settings is small. However, in most clinical and epidemiological studies, information from electronic health records (EHR), claims, pharmacies, etc., is needed.

To the best of our knowledge, there is a lack of studies describing the integration process of claims data, electronic health records, and prescriptions data into a single complete patient-centered view. In Estonia, three separate national electronic health databases store such information separately. These databases use different coding systems and structure which have hindered the co-use of these databases. To address these issues, we developed a reusable process to transform these three separate electronic health databases into a single coherent patient centric OMOP CDM. The current paper describes the transformation process, challenges faced, and solutions implemented.

## METHODS

### Data sources

Estonia is a small country in Northern-Eastern Europe with a population of 1.3 million consisting primarily of Estonians (70%). It is mandatory for all healthcare providers in Estonia to use three central operational health databases to enable easy data exchange between the institutions and interoperability. These databases cover clinical information from almost all healthcare settings (hospitals, specialists, family doctors, labs, pharmacies). Data could be potentially linked using personal identification codes provided to all residents. The main content and terminologies used in the databases are described in Table 1. There is some difference between the databases in the data coverage. Electronic health records (EHR) store data from all private and state-owned healthcare providers for insured and uninsured people, while health insurance claims include about 95% of the Estonian population who have public insurance[26]. Using the claims database is mandatory for reimbursement. All prescription drugs are prescribed digitally and stored in the corresponding database[27]. None of the health datasets is the primary source for death information (we were not allowed to use death registry data in this work), containing deaths related to healthcare services only (67%).

**Table 1.**
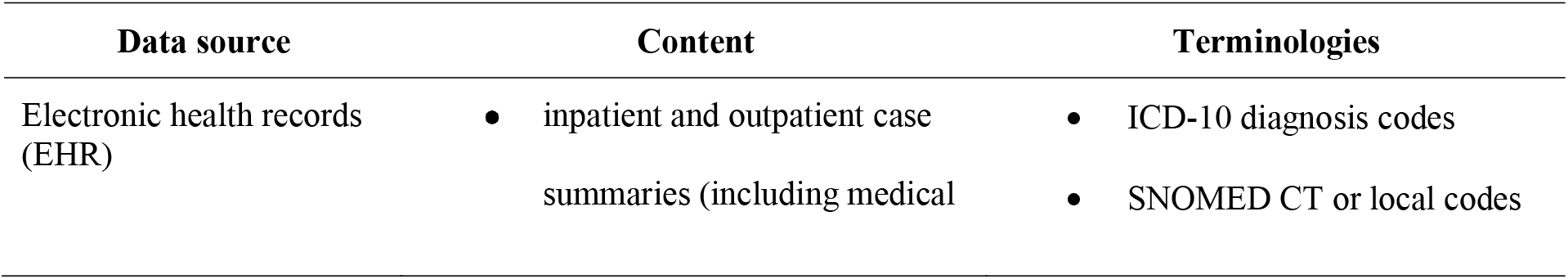
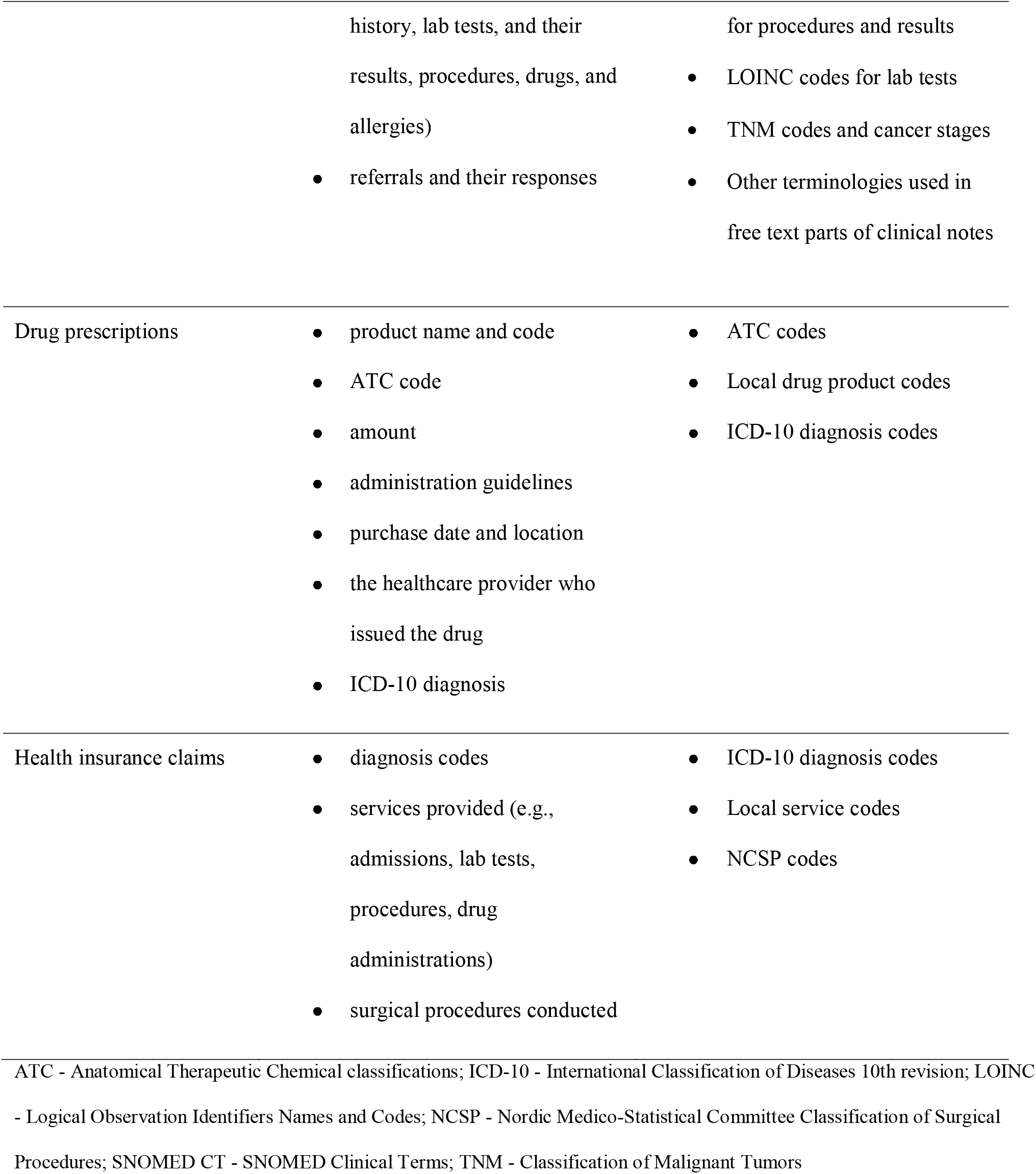
National health databases in Estonia.

Data from these three health databases complement each other; thus, linking adds additional value and improves the data quality. For example, health insurance claims provide information on which services (e.g., lab tests) have been provided, while further details of these services (e.g., the results of the tests) can be found in EHR. The drug prescription database does not contain information for over-the-counter medications where no prescription is needed (for example, paracetamol) or inpatient medications, but such information can be sometimes found in free text in EHR. At the same time, diagnoses, drugs, and procedures may be recorded in multiple source datasets, which adds complexity to the linking of health data. The drawback of EHR is its lower quality as the information is partially recorded in a semi-structured or free-text format (e.g., the weight or blood pressure of the patient is given in clinical notes) which is challenging to use in automated analysis. Also, case summaries in EHR contain only the most relevant information but may miss other tests or services conducted on a patient.

Carrying out an epidemiological study in Estonia requires obtaining approval from an ethics committee, followed by the collection of necessary data, typically from all three databases. To develop and demonstrate the transformation process of Estonian health data to OMOP CDM, we used a 10% random sample of the Estonian population (n = 150,824 patients) from 2012-2019 (Figure 1). For the sample, complete information from all three databases was extracted. This work was approved by the Estonian Bioethics and Human Research Council (EBIN, no. 1.1-12/653).

**Figure 1.**
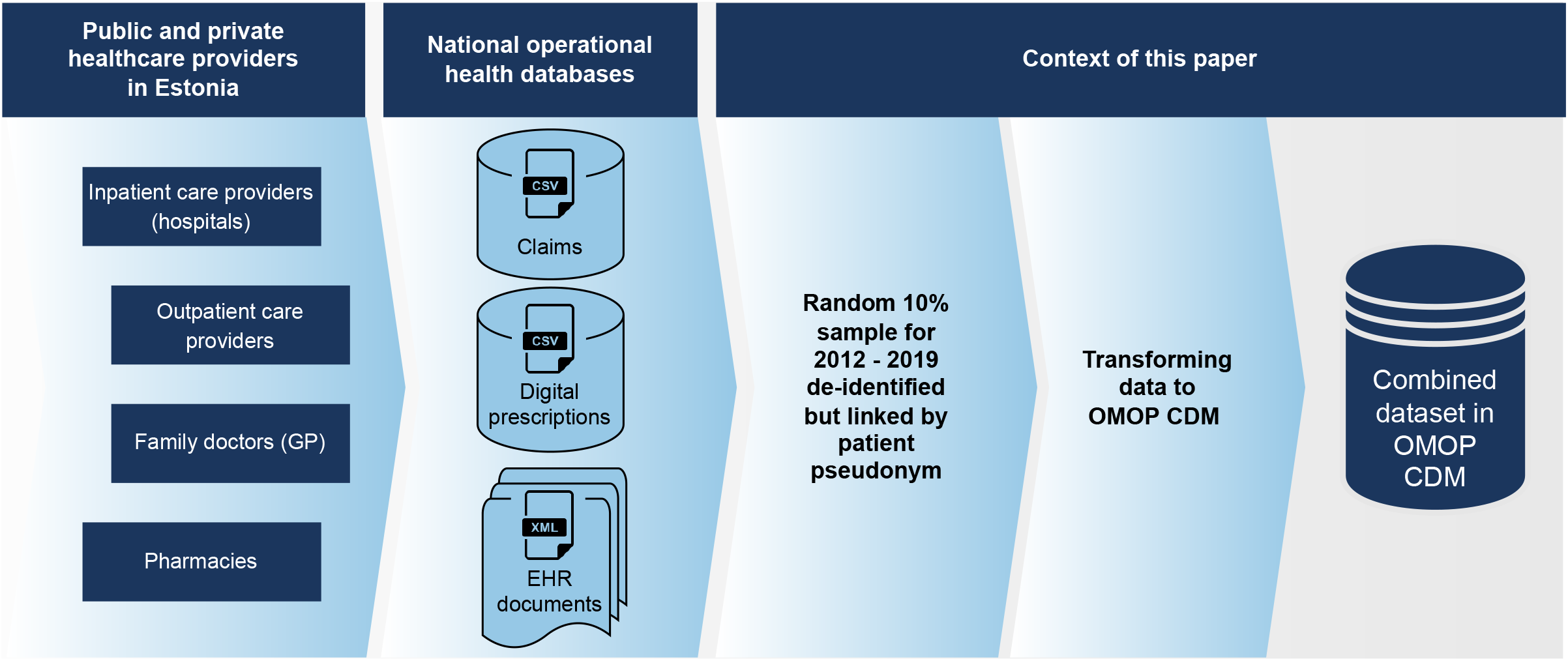
The data acquisition process of national health databases in Estonia and the context of this paper.

### Extract, transform, and load process

We transformed source data to the OMOP CDM version 5.3, which includes 15 clinical data tables for storing patient demographics and clinically relevant information. The transformation process had three main stages: first, creating the mapping between source and OMOP vocabularies; second, coming up with the technical implementation around the mapping process, starting with data extraction from source databases and ending with loading the data into the target database (extract, transform, load, ETL); and finally, validating the transformation results (Figure 2).

**Figure 2.**
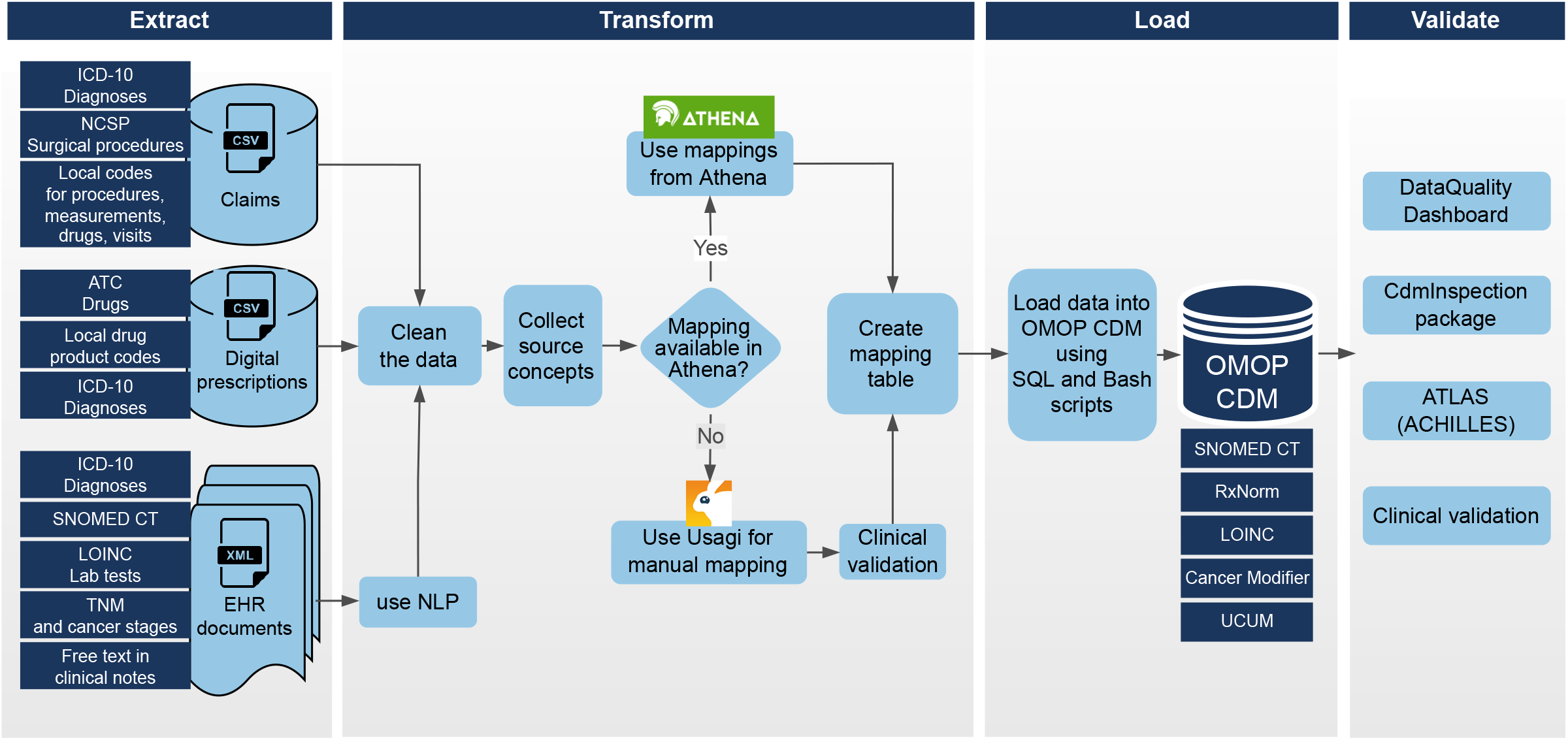
Overview of the data transformation process.

#### Mapping source vocabularies to OMOP vocabularies

Three source datasets (EHR, prescriptions, claims) used different terminologies to represent clinical events, and thus, they were all mapped to standard OMOP vocabularies. The standard vocabularies which are used in OMOP for medical information are SNOMED Clinical Terms (CT), RxNorm[28], Logical Observation Identifiers Names and Codes (LOINC), Unified Code for Units of Measure (UCUM), and OHDSI community-developed vocabularies (Cancer Modifier, OMOP Extension, and RxNorm Extension). As for non-standard vocabularies, two approaches were used. Where mappings between non-standard and standard vocabularies (e.g., from International Classification of Diseases 10th revision ICD-10 to SNOMED CT) already existed in the OHDSI vocabularies repository Athena[29], the existing mappings were used. Otherwise, manual mapping was performed, prioritizing the most frequent and essential terms for the planned research studies. For manual mapping, we used the Usagi[30] tool, an application provided by the OHDSI community to help create mappings between non-standard source concepts and standard concepts. Manual mappings were validated by medical specialists. Next, we give a more detailed overview of the mapping process based on different source codes in our databases.

The diagnosis codes in all three data sources were in ICD-10 format and were mapped to the OMOP vocabulary using the available mappings in Athena.

Lab tests from EHR were encoded using LOINC codes. As LOINC codes are standard in the OMOP vocabulary, no additional mapping was required. However, we needed additional mappings for lab test results. In cases where lab test results were presented as text (e.g., “negative” or “positive”), these were standardized, and the appropriate answer was determined based on the corresponding LOINC code. If a LOINC answer code was not present in the OMOP LOINC vocabulary or was determined to be invalid, a mapping to SNOMED CT was created.

To convert Anatomical Therapeutic Chemical classifications (ATC) codes to RxNorm, we used the standard ATC-to-RxNorm mapping available in Athena. However, as ATC codes provide information at the ingredient level and do not include information about strength or drug form, we also used Estonian-specific drug product codes to manually map source data to the RxNorm clinical drug level (ingredient, their strength, and form). If a drug product code could not be mapped, the ATC-to-RxNorm mapping was used instead.

The service codes are used on claims for administrative purposes. These are Estonian-specific codes, and the previous mapping to OMOP vocabularies was unavailable. These codes are divided into subgroups which include similar services, such as visits, procedures, surgeries, measurements without results, and drugs (e.g., specific cancer or hospital-given drugs). To map these codes to OMOP vocabularies, the codes were first translated to English, and then the Usagi tool was used to map these codes to valid SNOMED CT, RxNorm, or LOINC codes, depending on the nature of the source code.

Similarly to service codes, mappings for Nordic Medico-Statistical Committee Classification of Surgical Procedures (NCSP) codes and cancer-specific findings (TNM classification of malignant tumors, cancer stages, histopathology grades) were created using Usagi. The NCSP codes were mapped to standard SNOMED CT codes, and cancer-specific findings were mapped to the Cancer Modifier vocabulary.

If the source code could not be mapped to OMOP standard concept, the record was transferred to OMOP CDM with a concept identifier of “0” (meaning no matching concept) and placed in the appropriate target table based on the nature of the source concept. With this approach, the source concepts can still be used for defining studies within this dataset, however, such definitions likely will not work elsewhere.

#### Technical implementation

The ETL process was built using a combination of Bash scripts, SQL scripts, and manual comma-separated values (CSV) mapping files. The ETL process is shown in Figure 2. The process began with cleaning and validating the source data. From EHR documents, the necessary information was extracted from structured files of Extensible Markup Language (XML) and cleaned. As the format of the information is partially free-text or semi-structured in the EHR documents, natural language processing (NLP) methods were used to extract this information. The data from the three source datasets were combined into a single OMOP CDM database using developed pipelines and mappings. As there is no direct link between the same event in different data sources, no duplicate removal was performed. For example, if the same diagnosis code comes from an EHR document and claims, both are converted to OMOP CDM. Still, we determined the provenance of the information, whether it came from an EHR document, claim, or prescription data. The developed ETL process is reusable for other samples on these data sources.

#### Validation

The results of the ETL process were assessed using an open-source Achilles data characterization tool[31], DataQualityDashboard version 1.4.1[7], and the EHDEN CdmInspection tool[32]. Achilles data characterization tool[31] allows getting an overview of the converted data. DataQualityDasboard[7] runs over 3000 checks on conformance, completeness, and plausibility on the data transformed to OMOP CDM. CdmInspection tool[32] runs additional vocabulary (for example, top unmapped and mapped codes in different tables) and infrastructure checks compared to Achilles and DataQualityDashboard. Any errors, warnings, or issues found were addressed by revising the ETL code or mappings, and the process was repeated until all solvable errors were resolved.

## RESULTS

The transformation process for the OMOP CDM was developed using a sample of 10% of the Estonian population (n = 150,824 patients) with a similar age and gender distribution as the overall population (Figure 3). All persons have one observation period covering 1 Jan 2012 to 31 Dec 2019 except when the person was born later than 2012 or died before 2019. In these cases, the observation period was adjusted accordingly. In total, the sample dataset contained 4,970,022 EHR documents, 6,222,818 claims, and 9,289,527 digital prescriptions.

**Figure 3.**
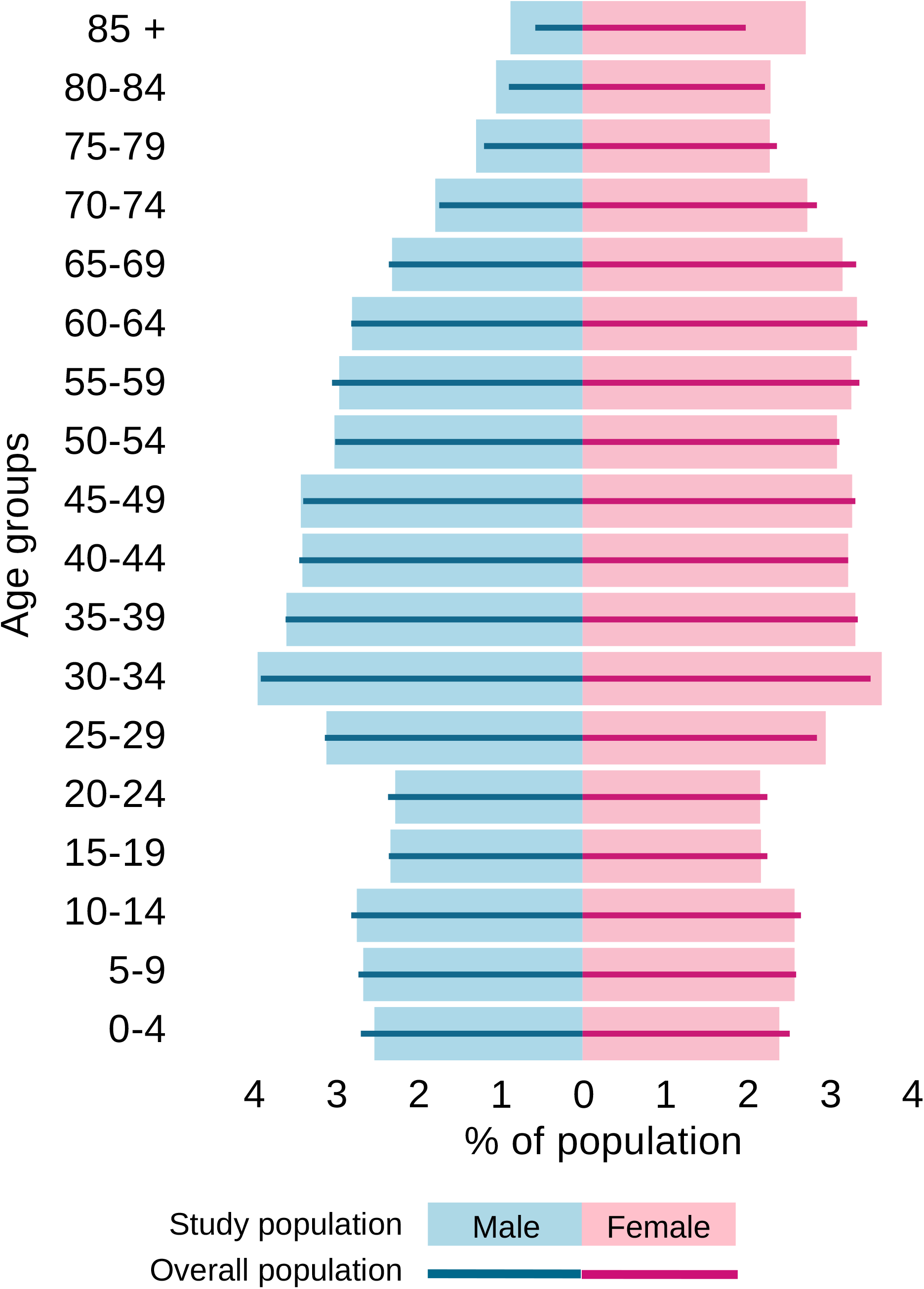
Population pyramids of an Estonian population in 2019 (lines) and the study sample (bars).

Out of 150,824 persons in the source data, we were able to transform 149,364 (99.0%) persons and their medical data to OMOP CDM. The remaining 1,460 persons did not have a birth year reported in any source datasets, which is a mandatory attribute in OMOP CDM. These patients had altogether 2,364 entries in source data. On the other hand, linking three datasets enabled us to determine the year of birth for 530 patients who were missing this information in some of the source datasets, and thus, we were able to include them in the target database.

The distribution of source data across target tables is shown in Table 2. Most populated target tables are for measurements (32,230,620 entries) and conditions (20,351,014). The highest percentage of entries mapped to OMOP standardized vocabularies are for tables visit_occurence and visit_detail (100%), condition_occurence (99.9%), drug_exposure (98.7%), observation (97.1%), and measurement (90.8%).

**Table 2.** Number of entries in OMOP CDM tables together with mapping rates

| OMOP CDM table | Total number of entries<br>in table | The number of entries<br>mapped to OMOP standard<br>concepts | Mapping rate |
| --- | --- | --- | --- |
| location | 1 | Not applicable | Not applicable |
| care_site | 1,820 | Not applicable | Not applicable |
| person | 149,364 | Not applicable | Not applicable |
| death | 8,277 | Not applicable | Not applicable |
| observation_period | 149,364 | Not applicable | Not applicable |
| visit_occurrence | 18,281,120 | 18,281,120 | 100.0% |
| visit_details | 48,002 | 48,002 | 100.0% |
| condition_occurrence | 20,351,014 | 20,333,065 | 99.9% |
| procedure_occurrence | 6,956,568 | 5,392,192 | 77.5% |
| drug_exposure | 7,945,992 | 7,842,231 | 98.7% |
| device_exposure | 77,842 | 47,296 | 60.8% |
| observation | 15,203,064 | 14,762,978 | 97.1% |
| measurement | 32,230,620 | 29,250,571 | 90.8% |

**Table 3** shows the mapping rate according to source vocabularies. Local service codes, ICD-10 codes, and LOINC codes were used the most in the data. For ICD-10 and LOINC codes, we covered almost all entries with standard concepts (100.0% and 99.3%, respectively). The coverage was also high for entries where local service codes were used (84.6%), although we only mapped 38.9% of source codes.

**Table 3.**
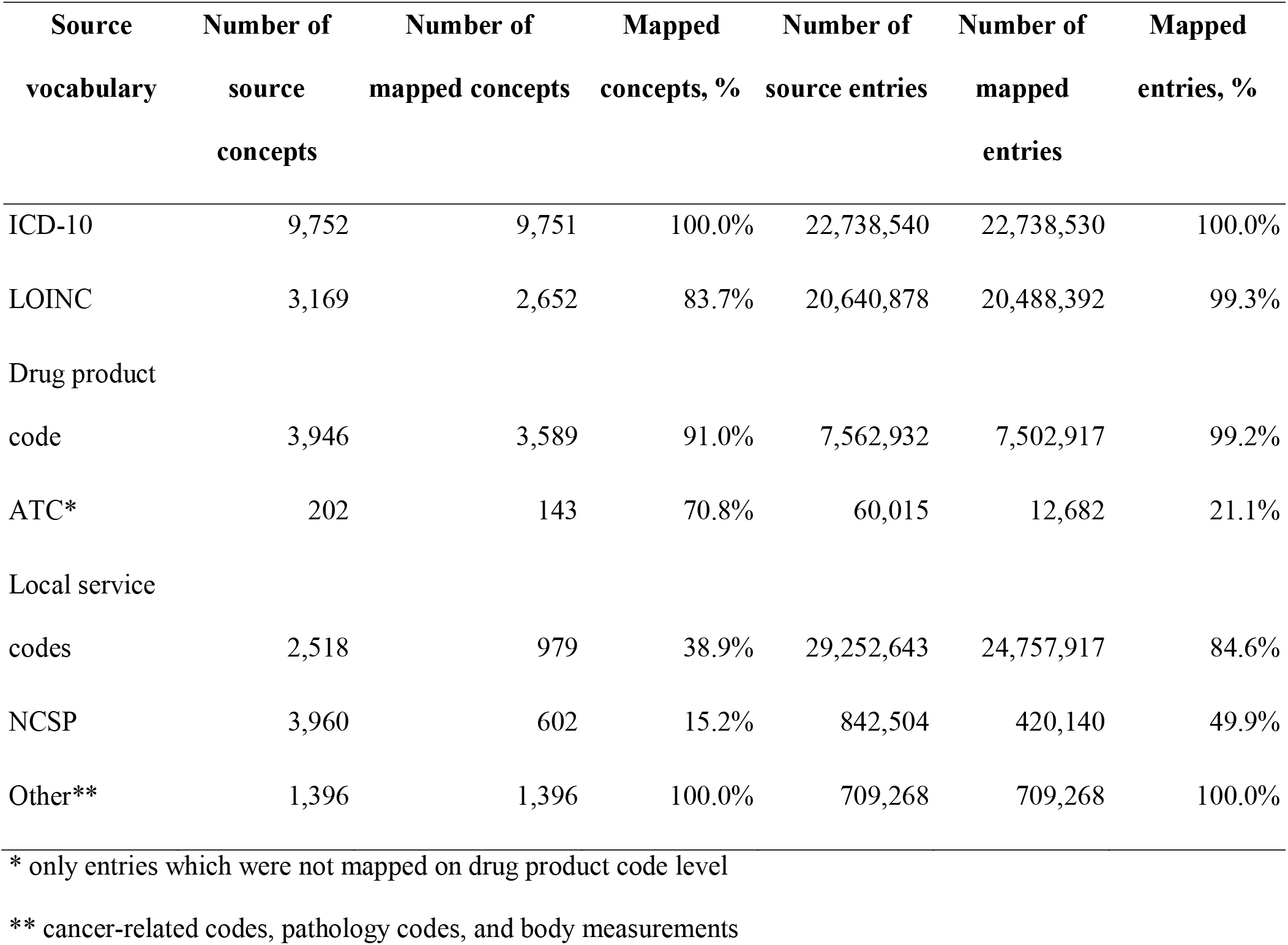
Mapped concepts and number of entries according to source vocabularies

The summary data of the output database can be found on a dedicated website[33]. The results of the DataQualityDashboard tool, which validates the plausibility, conformance, and completeness of the output dataset, are shown in Supplementary Table S1. Out of the 3,482 checks conducted, 3,431 passed, and 51 failed. The failed checks were evaluated individually, and it was determined that their failure was expected. Five plausibility checks failed because a gender-related clinical code was assigned to the wrong gender person. For example, four records with the concept of “primary malignant neoplasm of penis” were assigned to persons whose gender was female. For the completeness test, checks for five tables failed as the percentage of records where the standard concept value was “0” exceeded the threshold of 5%. This was due to the presence of unmapped records in the tables. Additionally, we had 41 checks failing due to the high proportion (>5%) of measurement values outside the range specified in DataQualityDashboard. Our investigation revealed that the values were within reasonable ranges (thus, no error in the data), and to our knowledge, the range windows have been removed in the next version of DataQualityDashboard.

## DISCUSSION

This paper describes the integration process of claims data, electronic health records, and prescriptions data into one complete patient-centered view. For a 10% random sample, three national health databases with complete records were linked and successfully transferred to OMOP CDM. Our experience shows that transferring health databases to OMOP CDM contains several challenges (Table 4). However, the outcome of the mapping and transformation process has a good quality and expands the possibilities for collaboration. To the best of our knowledge, this is one of the first papers of this kind and one of the largest by the proportion of the population of a country.

**Table 4.**
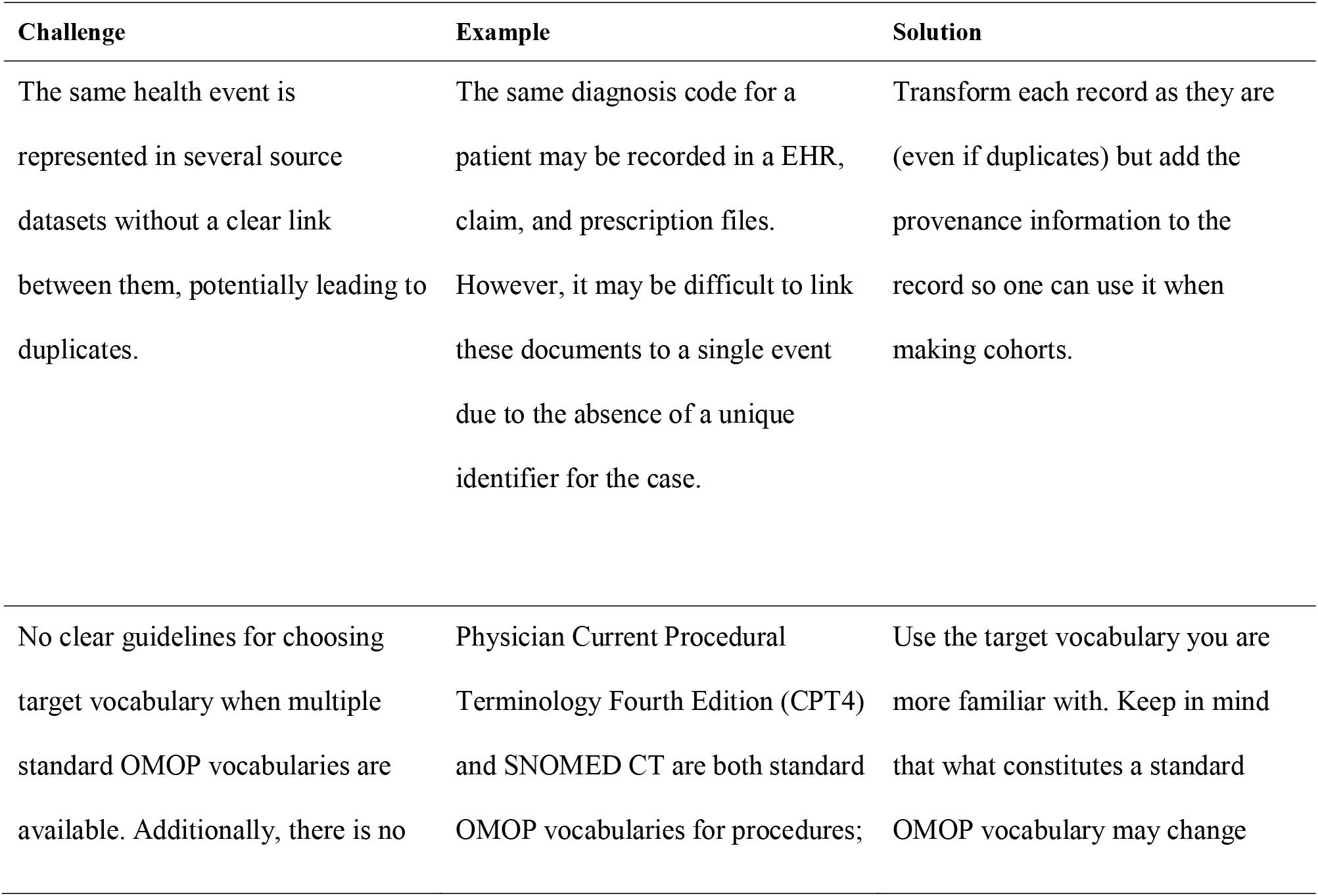
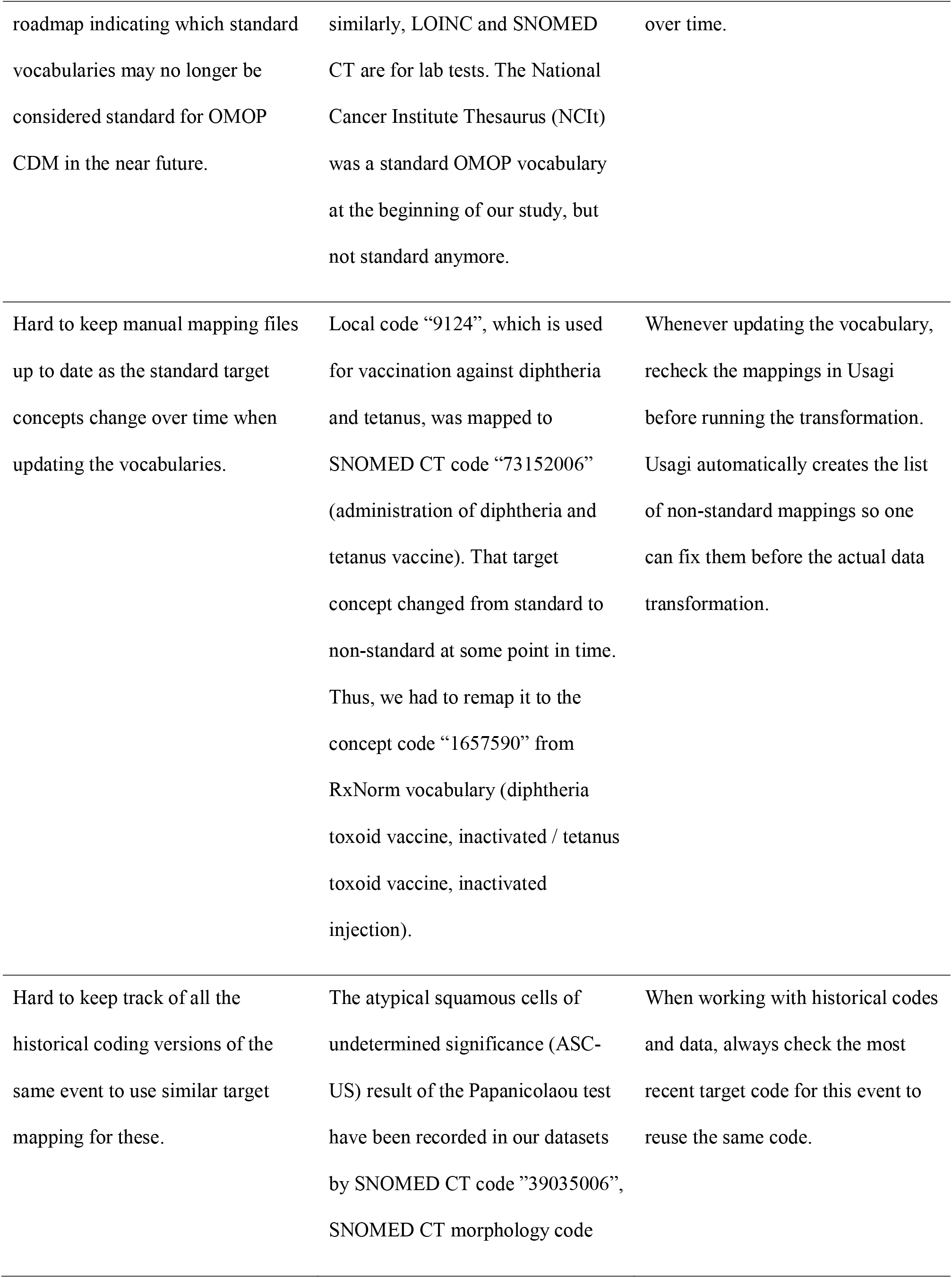
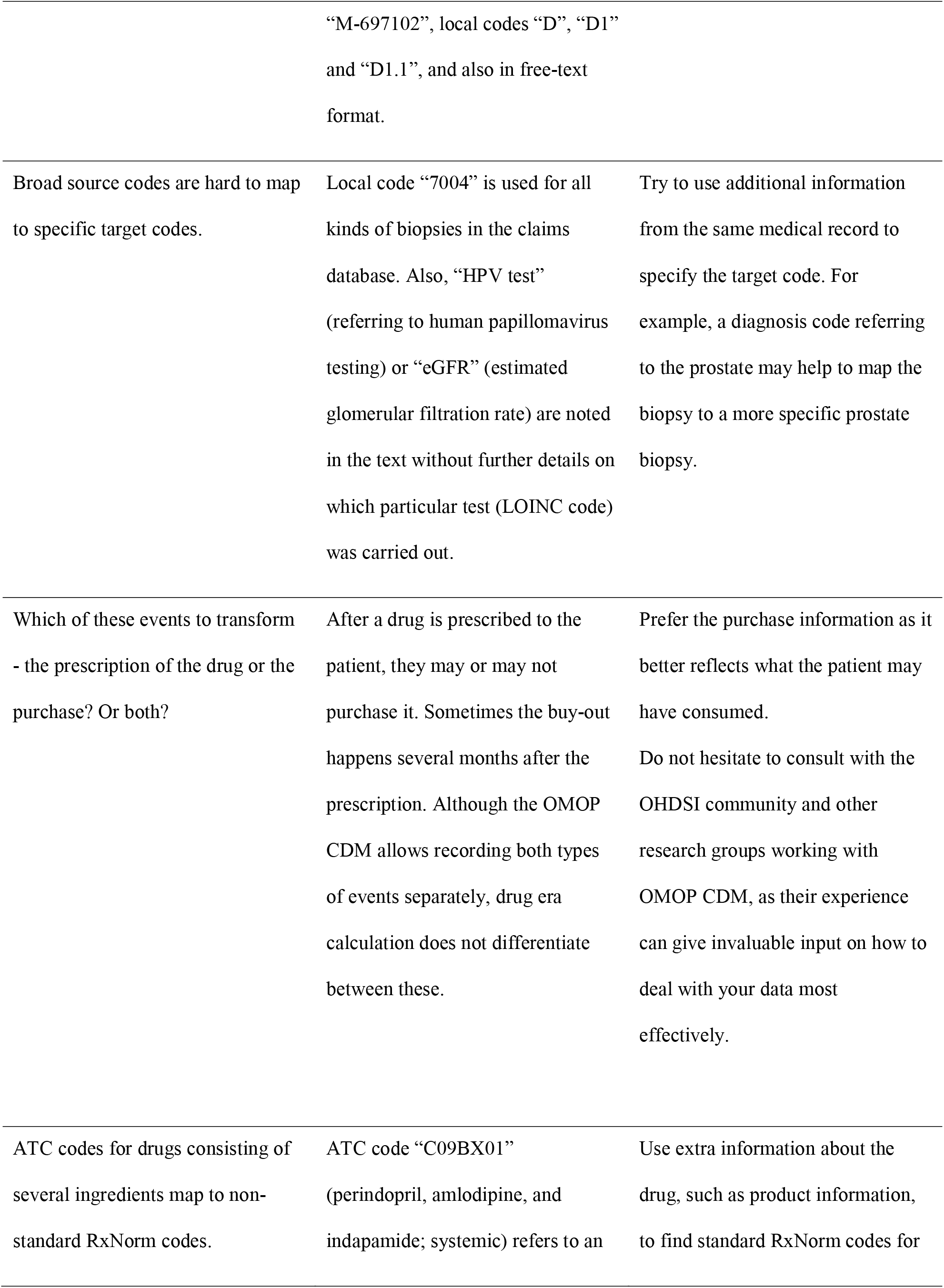
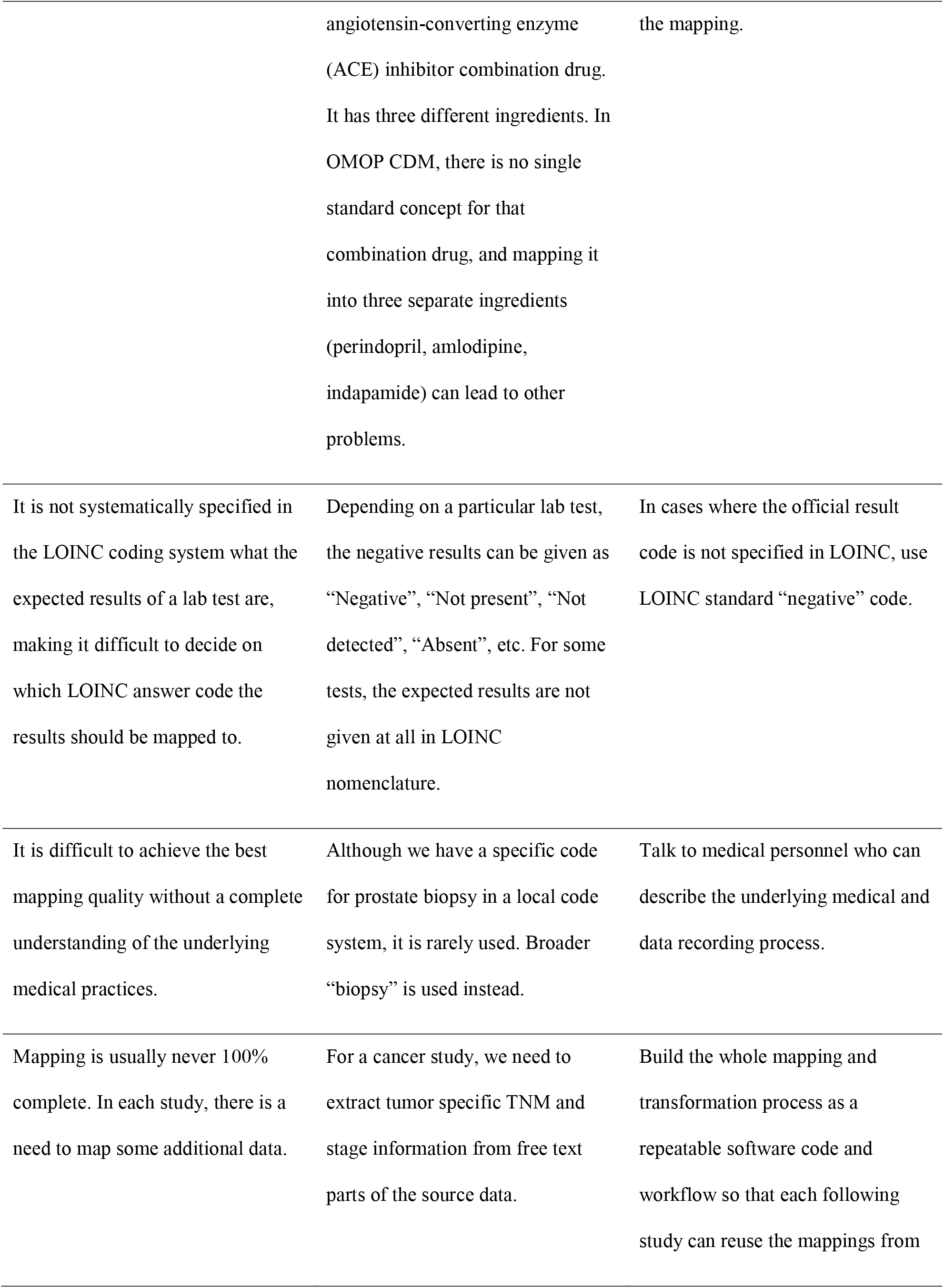
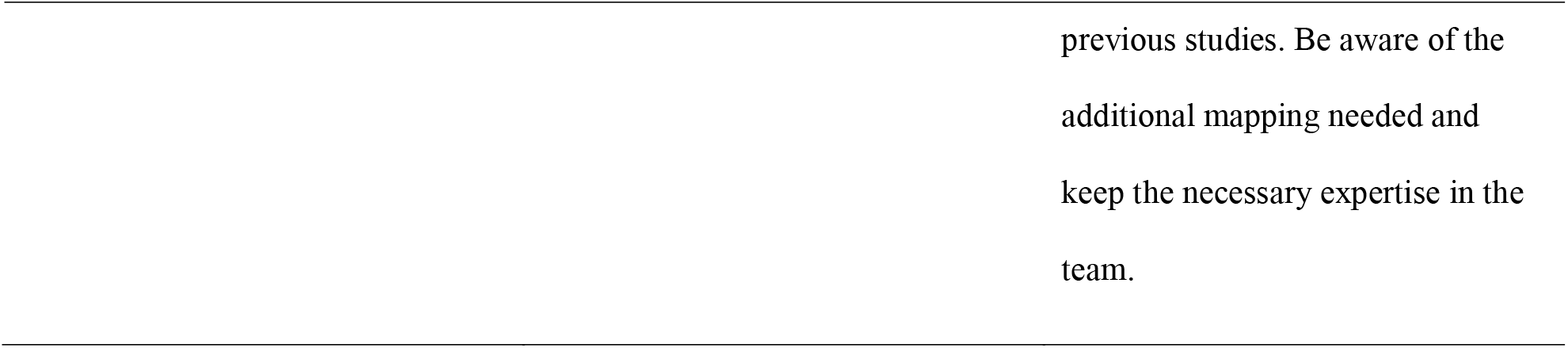
Main challenges and solutions of the current work

In total, we transformed over 100 million entries to standard concepts using standard OMOP vocabularies with the average mapping rate 95%. For conditions, observations, drugs, and measurements, the mapping rate was over 90%. In most cases, SNOMED CT was used as the target vocabulary. Similarly to others[11,13,16,21], we were not able to map all the records.

Previous studies have shown that one of the main difficulties during the transformation process is finding the relevant concepts[6,16,17,21,23]. This is also in line with our experience. In case source vocabulary was already considered standard vocabulary according to OMOP CDM or mapping between source and standard vocabulary was provided, we used that vocabulary. However, it is crucial to stay aware of the changes in standard vocabularies and to be prepared to update mappings continuously. There is also a possibility that standard vocabulary becomes non-standard over time. For instance, at the beginning of our mapping process, there were two standard vocabularies for cancer findings, but later, one of them was changed to non-standard by the OHDSI community, which created additional work. It would be helpful if the OHDSI community could provide clear recommendations for determining the most suitable target vocabularies for mapping to avoid potential issues in the future. When mappings to standard vocabularies are not present, there are, unfortunately, no established guidelines for determining which vocabulary should be used. We recommend selecting the target vocabulary that the user is most familiar with. We agree with the previously reported assertions that the most problematic is the mapping of local code systems[16,21,23] as ambiguity in source codes makes it difficult, if not impossible, to find appropriate target codes. This is the main reason why our mapping rate of local service codes is rather low (39%).

In addition to incomplete mapping, information loss during the transformation process can also occur due to data structure[23]. Our experience showed that integrating data from various healthcare settings can result in overlap, where the same health event or episode may be represented in multiple datasets. In case of overlapping events, a decision had to be made as to whether to treat these multiple instances of the same event (e.g., diagnosis) “as is” or to attempt to combine them into a single event in the OMOP CDM. After careful consideration, it was ultimately decided not to link the records and to use multiple instances of the event in the OMOP database. Several factors justified this decision. Firstly, the process of combining multiple records into a single event is complex and prone to error. Secondly, the transformation process would result in the loss of information about the exact source code and dataset, which may be necessary for quality control or specific studies. Finally, previous studies conducted using the OMOP CDM have not typically been concerned with the number of records or have required a minimum time interval between records, effectively addressing the issue of multiple close-time-range recordings of the same event[34,35].

Several actions can support and improve the transformation process. One of them is communication and collaboration with different experts and consortiums[36] It is mandatory to consult with the medical personnel to understand the clinical practice and map the codes correctly. Still, in our case, despite our efforts and the inclusion of medical experts, more than half of the local codes remained unmapped (61%). During the mapping process, we consulted with the OHDSI community about drug prescriptions data. According to the recommendations received, only drugs purchased from the pharmacy were mapped in our study. While information about prescribed medications may sometimes be more important than the fact of purchase, it was decided to exclude it to minimize confusion in the execution phase of future studies. In addition, participation in international projects and consortiums can provide insight into any additional data requirements, existing problems, and necessary mappings for specific studies. By having one modular codebase, this knowledge will accumulate and can be built upon in subsequent studies.

When planning a data transformation to OMOP CDM, it should be considered that transforming one or multiple linked datasets to a common data model cannot be taken as a one-time project but rather a continuous and iterative process requiring dedicated personnel, tools, and experience. This recommendation was previously highlighted by Candore et al.[36] as well. For example, conducting a study on the created dataset can reveal issues in the data that were missed during the transformation. This means that some transformation steps must be repeated to improve the data quality. Due to the continuous and iterative nature of the transformation process, our experience highlights that it is essential to have the entire process as a version-controlled software code that can be reused, including any mappings created in previous studies. This will eventually lead to a gradual improvement in the quality of the dataset.

Despite our efforts, our work has some limitations that must be considered. Firstly, the described transformation pipeline with the created mappings is directly applicable to Estonian national datasets only. Secondly, the created dataset cannot be made publicly available. Also, as the dataset includes data from 2012–2019, the current observation period can be too short for some studies. At the same time, the age and gender distribution of the sample follow the whole population; thus, the dataset can be considered representative. To date, the work is still ongoing to improve the quality of the dataset and extract as much important information as possible. For example, the mapping coverage for NSCP classification of surgical procedures or device exposures (Table 2 and Table 3) is currently modest, as these have not been the focus of our current research. Moreover, there is still a large amount of information stored as free text, which is being gradually extracted.

As a result of the transformation process, we have created a rare example of Northern-Eastern European datasets mapped to OMOP CDM containing data from the 10% of Estonian population and almost all healthcare settings. To our knowledge, the harmonization and integration of these three national datasets have been a unique and innovative effort, even for a large OHDSI community. The usefulness of the dataset has been demonstrated through its application in various national and international studies and projects for generating evidence. For example, in Estonia, we have performed a study investigating the presence of HPV virus types and cervical cytology grades[37] and analyzed how artificial intelligence could be applied to health data for public service[38]. In addition, developing a tool for analyzing health event trajectories in any OMOP dataset[39] or participating in the study-a-thon of a project to harness big data in prostate cancer research[40] would not have been possible without the transformation process. We have validated the data linkage and the above describe repeatable approach in the PIONEER study, where the cohort of patients with newly diagnosed prostate cancer had an inclusion criterion requiring both a diagnosis and biopsy to be recorded[41]. Using only EHR or prescription data would have yielded zero patients in Estonia while using only claims data would have yielded 235 patients. However, combining these two datasets resulted in a person count of 635 patients. The repeatable transformation scripts have been reused with only minor adjustments on independent new cohorts on the prescriptions of 110,000 asthma patients, insurance claims and prescriptions of 400,000 COVID-19 patients with controls, and on all the medical data of more than 200,000 gene donors of Estonian Biobank. This all has significantly contributed to the efficient use of real-world data.

## CONCLUSION

This paper contributes to the broader use of real-world data. We have described our approach to link three central health databases in Estonia and successfully demonstrated transferring 10% of the data to OMOP CDM. The methods described can be applied to any future study using Estonian health data, and could potentially be used to convert the entire population’s health data to OMOP CDM. Additionally, these principles can be applied beyond Estonia. Despite the challenges faced during the transformation process, our experience shows that OMOP CDM can be effectively used for healthcare data and that the transformation can increase the opportunities for health data analysis and collaboration. Our work helps future researchers to transform linked databases into OMOP CDM more efficiently, ultimately leading to better real-world evidence.

## Data Availability

The data underlying this article cannot be shared publicly for the privacy of individuals that participated in the study. The data was obtained from national health databases and can be requested via Estonian Bioethics and Human Research Council.

## ACKNOWLEDGEMENTS

The data processing described in the article was carried out at the High Performance Computing Center of the University of Tartu.

## SUPPLEMENTARY MATERIAL

**Supplementary Table S1 -** DataQualityDashboard results of the data transformed to OMOP CDM

