## Supplementary figures and images for "Transforming Estonian health data to the Observational Medical Outcomes Partnership (OMOP) Common Data Model: lessons learned"

### Supplementary Table S1

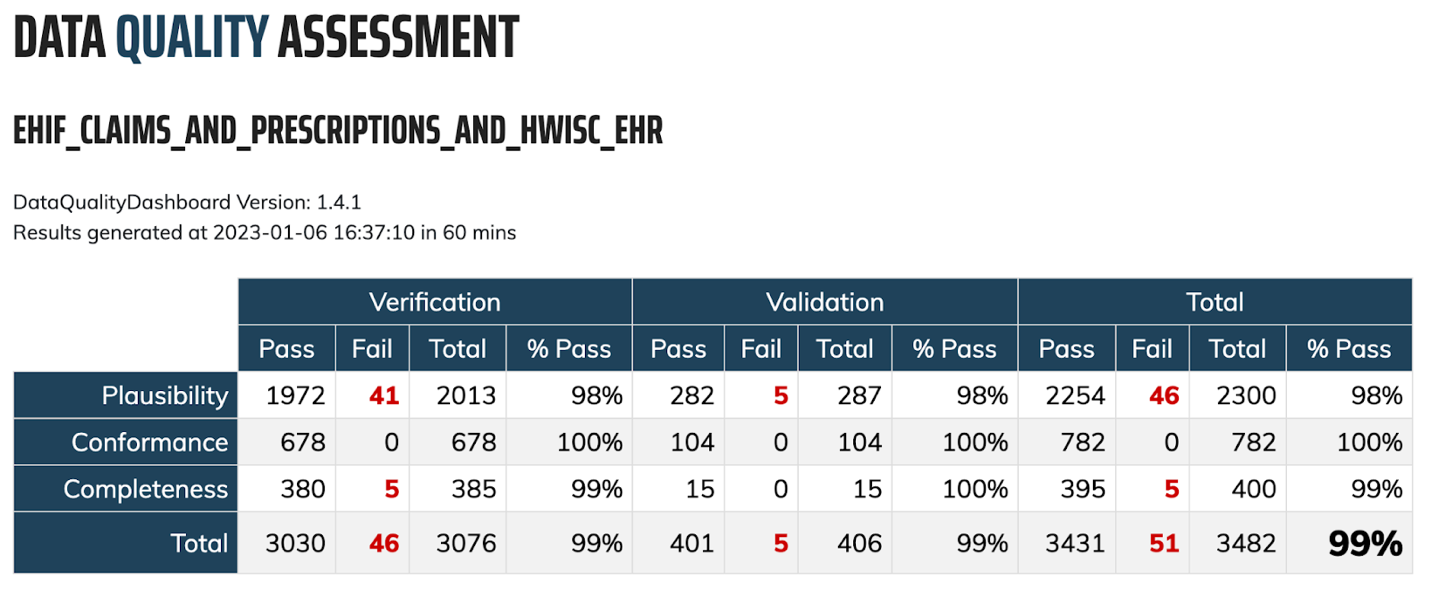

**Supplementary Table S1.** DataQualityDashboard results of the data transformed to OMOP CDM
